# Dietary supplements during the COVID-19 pandemic: insights from 1.4M users of the COVID Symptom Study app - a longitudinal app-based community survey

**DOI:** 10.1101/2020.11.27.20239087

**Authors:** Panayiotis Louca, Benjamin Murray, Kerstin Klaser, Mark S Graham, Mohsen Mazidi, Emily R Leeming, Ellen Thompson, Ruth Bowyer, David A Drew, Long H Nguyen, Jordi Merino, Maria Gomez, Olatz Mompeo, Ricardo Costeira, Carole H Sudre, Rachel Gibson, Claire J Steves, Jonathan Wolf, Paul W Franks, Sebastien Ourselin, Andrew T Chan, Sarah E Berry, Ana M Valdes, Philip C Calder, Tim D Spector, Cristina Menni

## Abstract

**Objectives:** Dietary supplements may provide nutrients of relevance to ameliorate SARS-CoV-2 infection, although scientific evidence to support a role is lacking. We investigate whether the regular use of dietary supplements can reduce the risk of testing positive for SARS-CoV-2 infection in around 1.4M users of the COVID Symptom Study App who completed a supplement use questionnaire.

**Design:** Longitudinal app-based community survey and nested case control study.

**Setting:** Subscribers to an app that was launched to enable self-reported information related to SARS-CoV-2 infection for use in the general population in three countries.

**Main Exposure:** Self-reported regular dietary supplement usage since the beginning of the pandemic.

**Main Outcome Measures:** SARS-CoV-2 infection confirmed by viral RNA polymerase chain reaction test (RT-PCR) or serology test. A secondary outcome was new-onset anosmia.

**Results:** In an analysis including 327,720 UK participants, the use of probiotics, omega-3 fatty acids, multivitamins or vitamin D was associated with a lower risk of SARS-CoV-2 infection by 14%(95%CI: [8%,19%]), 12%(95%CI: [8%,16%]), 13%(95%CI: [10%,16%]) and 9%(95%CI: [6%,12%]), respectively, after adjusting for potential confounders. No effect was observed for vitamin C, zinc or garlic supplements. When analyses were stratified by sex, age and body mass index (BMI), the protective associations for probiotics, omega-3 fatty acids, multivitamins and vitamin D were observed in females across all ages and BMI groups, but were not seen in men. The same overall pattern of association was observed in both the US and Swedish cohorts. Results were further confirmed in a sub-analysis of 993,365 regular app users who were not tested for SARS-CoV-2 with cases (n= 126,556) defined as those with new onset anosmia (the strongest COVID-19 predictor).

**Conclusion:** We observed a modest but significant association between use of probiotics, omega-3 fatty acid, multivitamin or vitamin D supplements and lower risk of testing positive for SARS-CoV-2 in women. No clear benefits for men were observed nor any effect of vitamin C, garlic or zinc for men or women. Randomised controlled trials of selected supplements would be required to confirm these observational findings before any therapeutic recommendations can be made.

## Introduction

A number of micronutrients, including vitamins C and D and zinc, have been shown to play key roles in supporting immune function [1,2] and in reducing risk of respiratory infection [2,3]. These nutrients can be obtained from the diet but are also available as dietary supplements either alone or as part of multivitamin or multinutrient mixtures. There are many other dietary supplements available including omega-3 fatty acids (“fish oil”), probiotics and plant isolates like garlic [4]. The use of specific dietary supplements in both prevention and acute treatment of infection with SARS-CoV-2 has been promoted since the beginning of the current coronavirus pandemic [5]. The UK supplement market increased by 19.5% in the period leading up to the national “lockdown” in early March 2020 [6], with a 110% rise in sales of vitamin C and a 93% rise in sales of multivitamin supplements [6]. Likewise, zinc supplement sales increased by 415% over the 7-day period ending 8^th^ March, at the height of COVID-19 concern in the US [5].

A biologically plausible role exists for the use of certain dietary supplements [1]. For example, vitamin D has been suggested to reduce SARS-CoV-2 transmission by enhancing antiviral immunity and to reduce mortality mitigating the cytokine storm linked with severe COVID-19 [7,8]. Moreover, zinc also supports the function of the immune system [9] and may have specific antiviral effects [10]. However, robust evidence to support a role for dietary supplements in preventing infection with SARS-CoV-2 is not available [11]. Any such evidence would need to take into account factors such as socioeconomic status, ethnicity, and occupational exposure to the virus as well as the requirement of a large sample size and a clear confirmation of infection.

By using data from the COVID Symptom Study App [12] on 4,544,666 users in the UK, we tested the hypothesis that use of dietary supplements would be associated with a lower risk of testing positive for SARS-CoV-2. We initially examined whether supplement use was associated with SARS-CoV-2 infection among 327,720 UK participants who reported having been tested for SARS-CoV-2 using a reverse transcriptase polymerase chain reaction (RT-PCR)- or serology-based test. Next, we used data from 45,757 US and 27,373 Swedish users of the app who also reported tests for SARS-CoV-2 infection to replicate UK findings. Finally, in an independent sub-analysis we examined the extent to which supplement use was associated with self-reported anosmia, the single strongest predictor of COVID-19 [12,13], in 993,365 regular app users who were not tested for SARS-CoV-2.

## Methods

### Study setting and participants

The COVID Symptom Study app was developed by health data company Zoe Global Ltd with input from King’s College London, the Massachusetts General Hospital, Lund University, Sweden and Uppsala University, Sweden. It was launched in the UK on Tuesday the 24^th^ March 2020 and in the US on Sunday the 29^th^ March 2020 as previously described [12,14]. Once translated, the app was launched in Sweden on 29^th^ April 2020. The app enabled self-reported information related to SARS-CoV-2 infection to be captured. On first use, the app recorded self-reported location, age, and core health risk factors. With continued use, participants provided daily updates on symptoms, health care visits, SARS-CoV-2 test results, and if they were self-quarantining or seeking health care, including the level of intervention and related outcomes. Individuals without apparent symptoms were also encouraged to use the app. Via the app, data on regular use (more than 3 times a week for at least three months) was collected from June 2020 (See **Table S1** for list of questions).

These, include use of probiotics, garlic, omega-3 fatty acids (“fish oils”), multivitamins, vitamin D, vitamin C or zinc. Supplement use was recorded as yes/no. Through direct updates, new or modified questions were added in real-time to capture data to test emerging hypotheses about COVID-19 symptoms and treatments. A subset of 234,271 UK app users also completed an online short form food frequency questionnaire, from which a validated Diet Quality Score was generated. This work was conducted using the Short Form FFQ tool developed by Cleghorn and collaborators and listed in the Nutritools (www.nutritools.org) library[15].

### Assessment of Exposure

Information on the exposure was collected via the app. We included a subset of individuals who reported being tested for SARS-CoV-2 infection using RT-PCR or serology and answered the supplements questionnaire.

### Ascertainment of outcomes

Participants were asked if they had been tested for COVID-19 and the results (none, negative, pending, or positive). Our primary outcome was a report of a positive COVID-19 test. Follow-up started when participants first reported on the COVID Symptom Study app and continued until a report of a positive COVID-19 test or the time of last data entry, whichever occurred first. We also included an independent subset of 993,365 regular app users (reporting at least 30 times since creating their profile) who were not tested for SARS-CoV-2. Cases were defined as those reporting new onset anosmia, while non-cases were app users free from symptoms throughout the study.

### Ascertainment of covariates

Covariates including age, sex, BMI, smoking, ethnicity, healthcare worker status, and presence of comorbidities (i.e. cancer, diabetes, eczema, heart disease, lung disease, kidney disease and hay fever) were self-reported via the app. The app also facilitated the index of multiple deprivation (IMD) to be generated from the relevant government websites GB [16], Scotland [17], Wales [18], with the most recent IMD available at the time of analysis used. The IMD was then categorised into quintiles within-population, where 1 is the least deprived and 5 is the most deprived. A subset of UK participants undertook a Leeds short-form food frequency questionnaire, from which a dietary quality index was derived.

### Ethics

Ethical approval for use of the app for research purposes in the UK was obtained from King’s College London Ethics Committee (review reference LRS-19/20-18210) and all users provided consent for non-commercial use. The US protocol was approved by the Partners Human Research Committee (protocol 2020P000909) [19]. The Swedish protocol was approved by the Swedish Ethical Review Authority.

### Data sharing

Anonymised research data are shared with third parties via the centre for Health Data Research UK (HDRUK.ac.uk). US investigators are encouraged to coordinate data requests through the COPE Consortium (www.monganinstitute.org/cope-consortium). Data updates can be found on https://covid.joinzoe.com

### Statistical analysis

We studied 372,720 UK app users (aged 16-90 years). Of these, 23,521 individuals tested positive for SARS-CoV-2 and 349,199 tested negative. Multivariate logistic regression adjusting for age, sex, body mass index (BMI) and health status at sign-up was applied to investigate the association between supplement use and testing positive for SARS-CoV-2. We then repeated the analyses (i) adjusting for age, sex, BMI, comorbidities (including type-2 diabetes, cancer, asthma, heart disease, eczema, hay fever, kidney disease and lung disease), index of multiple deprivation, smoking, ethnicity, health worker/carer status and diet quality and (ii) stratifying by sex, age group (< 40, 40-60, > 60 years) and BMI categories (normal weight, overweight, obese/morbidly obese).

Replication was conducted in two independent datasets including 45,757 US and 27,373 Swedish (SE) app users (**Figure 2**).

Finally, in a sub-analysis, multivariate logistic regression adjusting for age, sex, BMI, country and health status at sign-up was applied to validate the association between supplement use and having COVID-19 defined as new onset anosmia in 92,578 anosmia cases and 900,787 non-cases.

All P values presented were two-sided, with statistical significance determined by the Bonferroni corrected threshold of significance (P=0.05/7=0.007). Statistical analysis was performed using Stata v12 and ExeTera, a Python library developed at KCL to clean and process the raw dataset [20].

### Patient and public involvement

No patients were directly involved in designing the research question or in conducting the research. No patients were asked for advice on interpretation or writing up the results. There are no plans to involve patients or relevant patient community in dissemination at this moment.

## Results

The demographic characteristics of the study population are presented in **Table 1**. Briefly, our discovery cohort included 372,720 UK app users who reported having had an RT-PCR-based or serology test for SARS-CoV-2 and who completed the app-based dietary supplement questionnaire. The study sample was predominantly female (66.8%) and more than 50% were overweight (BMI(SD) = 26.8(5.6) kg/m^2^).

**Table 1.** Demographic Characteristics of the Study Population.

|  | UK (n=372,720) |  | US (n=45,757) |  | SE (n=27,373) |  | Validation analysis (n=993,365) |  |
| --- | --- | --- | --- | --- | --- | --- | --- | --- |
|  | Supplement users |  | Supplement users |  | Supplement users |  | COVID-19 symptom - Anosmia |  |
|  | YES | NO | YES | NO | YES | NO | YES | NO |
| **N (%)** | 175,652<br>(47.1%) | 197,068<br>(52.9%) | 32,314<br>(70.6%) | 13,443<br>(29.4%) | 13,422<br>(49%) | 13,951<br>(51%) | 92,578<br>(9.3%) | 900,787<br>(90.7%) |
| **SARS-CoV-2 positive, N (%)** | 10,508<br>(6%) | 13,013<br>(6.6%) | 2,002<br>(6.2%) | 1,211<br>(9%) | 1,806<br>(13.5%) | 2,206<br>(15.8%) | - | - |
| **Females, N(%)** | 123,462<br>(70.3%) | 125,651<br>(63.8%) | 22,817<br>(70.6%) | 8,210<br>(61.1%) | 9,694<br>(72.2%) | 9,088<br>(65.1%) | 54,363<br>(58.7%) | 522,473<br>(58%) |
| **White, N(%)** | 163,479<br>(93.1%) | 188,030<br>(95.4%) | 28,143<br>(87.1%) | 11,757<br>(87.5%) | 13,411<br>(99.9%) | 13,943<br>(99.9%) | 83,153<br>(89.8%) | 821,610<br>(91.2%) |
| **Current smoker N(%)** | 6,004<br>(3.4%) | 10,773<br>(5.5%) | 1,252<br>(3.9%) | 924<br>(6.9%) | 735<br>(5.5%) | 795<br>(5.7%) | 5,103<br>(5.5%) | 37,050<br>(4.1%) |
|  | *Mean (SD)* | *Mean (SD)* | *Mean (SD)* | *Mean (SD)* | *Mean (SD)* | *Mean (SD)* | *Mean (SD)* | *Mean (SD)* |
| **Age, yrs** | 49.57 (14.2) | 46.26 (14.4) | 56.24 (15.2) | 47.8<br>(16.0) | 49.0<br>(13.0) | 46.63<br>(12.9) | 49.26<br>(16.5) | 53.3<br>(15.7) |
| **BMI, kg/m<sup>2</sup>** | 26.59 (5.6) | 27.04 (5.7) | 27.27 (5.9) | 27.21 (6) | 26.1 (4.8) | 26 (4.7) | 26.5 (5.4) | 26.49 (5.3) |
| **IMD, median (IQR)** | 7 (5) | 6 (5) | - | - | - | - | - | - |
| **DQI, median (IQR)** | 11 (3) | 11 (3) | - | - | - | - | - | - |
| **Supplement use** |  |  |  |  |  |  |  |  |
|  | *n*<br>(%) | *n*<br>(%) | *n*<br>(%) | *n*<br>(%) | *n*<br>(%) | *n*<br>(%) | *n*<br>(%) | *n*<br>(%) |
| **Omega-3** | 39,263<br>(22.4%) | - | 8,663<br>(26.8%) | - | 3,039<br>(22.6%) | - | 10,190<br>(11%) | 113,169<br>(12.6%) |
| **Probiotics** | 20,449<br>(11.6%) | - | 7,268<br>(22.5%) | - | 1,715<br>(12.8%) | - | 4,484<br>(4.8%) | 46,411<br>(5.2%) |
| **Garlic** | 7,235<br>(4.1%) | - | 1,137<br>(3.5%) | - | 941<br>(7%) | - | 1,931<br>(2.1%) | 18,868<br>(2.1%) |
| **Multivitamins** | 77,034<br>(43.9%) | - | 18,843<br>(58.3%) | - | 5,496<br>(41%) | - | 17,998<br>(19.4%) | 174,641<br>(19.4%) |
| **Vitamin D** | 86,190<br>(49.1%) | - | 19,444<br>(60.2%) | - | 6,722<br>(50.1%) | - | 18,436<br>(19.9%) | 194,828<br>(21.6%) |
| **Vitamin C** | 46,755<br>(26.6%) | - | 10,136<br>(31.4%) | - | 4,045<br>(30.1%) | - | 10,407<br>(11.2%) | 104,238<br>(11.6%) |
| **Zinc** | 21,776<br>(12.4%) | - | 4,330<br>(13.4%) | - | 2,394<br>(17.8%) | - | 4,310<br>(4.7%) | 45,489<br>(5%) |
| **Pre-existing medical conditions** |  |  |  |  |  |  |  |  |
| **Type-2 diabetes** | 5,047<br>(2.9%) | 4,842<br>(2.5%) | 1,787<br>(5.5%) | 437<br>(3.3%) | 331<br>(2.5%%) | 251<br>(1.8%%) | 2,747<br>(3%) | 32,234<br>(3.6%) |
| **Cancer** | 2,178<br>(1.2%) | 1,902<br>(1%) | 977<br>(3%) | 217<br>(1.6%) | 157<br>(1.2%) | 130<br>(0.9%) | 1,005<br>(1.1%) | 10,791<br>(1.2%) |
| **Asthma** | 27,146<br>(15.5%) | 26,020<br>(13.2%) | 5,015<br>(15.5%) | 1,788<br>(13.3%) | 2,039<br>(15.2%) | 1,673<br>(12%) | 10,880<br>(11.8%) | 102,366<br>(11.4%) |
| **Heart disease** | 5,708<br>(3.3%) | 5,278<br>(2.7%) | 2,388<br>(7.4%) | 590<br>(4.4%) | 654<br>(4.9%) | 592<br>(4.2%) | 3,154<br>(3.4%) | 36,266<br>(4%) |
| **Eczema** | 22,896<br>(13%) | 23,454<br>(11.9%) | 3,423<br>(10.6%) | 1,346<br>(10%) | 1,768<br>(13.2%) | 1,779<br>(12.8%) | 9,534<br>(10.3%) | 91,426<br>(10.2%) |
| **Hay fever** | 79,024<br>(45%) | 80,114<br>(40.7%) | 17,000<br>(52.6%) | 6,256<br>(46.5%) | 5,497<br>(41%) | 5,256<br>(37.7%) | 35,209<br>(38%) | 342,863<br>(38.1) |
| **Kidney disease** | 1,702<br>(1%) | 1,542<br>(0.8%) | 630<br>(2%) | 143<br>(1.1%) | 117<br>(0.9%) | 102<br>(0.7%) | 722<br>(0.8%) | 7,737<br>(0.9%) |
| **Lung disease** | 24,651<br>(14%) | 22,924<br>(11.6%) | 4,331<br>(13.4%) | 1,329<br>(9.9%) | 1,894<br>(14.1%) | 1,488<br>(10.1%) | 9,090<br>(9.8%) | 96,888<br>(10.8%) |

IMD, index of multiple deprivation; BMI, body mass index; DQI, Diet Quality Score; IQR, Inter-quartile range; SE, Sweden. Self-reported comorbidities were not asked at sign-up and therefore not reported by all subjects

As shown in **Table 1**, out of the 372,720 UK app users, 175,652 self-reported using supplements regularly since the beginning of the pandemic, while 197,068 self-reported they were not.

In the UK cohort, regular supplementation with (i) multivitamins was associated with lower risk of testing positive for SARS-CoV-2 by 13% (OR[95%CI]= 0.87[0.84,0.90], P=1.62×10^−14^), (ii) vitamin D was associated with lower risk by 9% (OR[95%CI]= 0.91[0.88,0.94], P=2.07×10^−8^); (iii) probiotics was associated with lower risk by 14% OR[95%CI]=0.86[0.81,0.92], P=1.99×10^−6^); and (iv) omega-3 fatty acids was associated with lower risk by 12% (OR[95%CI]= 0.88[0.84,0.92], P=5.8×10^−8^), after adjusting for age, sex, BMI, sign-up health status and multiple testing (**Figure 1**). There were no significant associations for supplementation with zinc, vitamin C or garlic (**Figure 1**). To account for a potential healthy user bias, we did a sensitivity analysis further adjusting for ethnicity, comorbidities, smoking, index of multiple deprivation, health worker/carer status and diet quality and results were consistent with the previous analysis (**Figure 1**) though the effect of probiotics appeared weaker.

**Figure 1.**
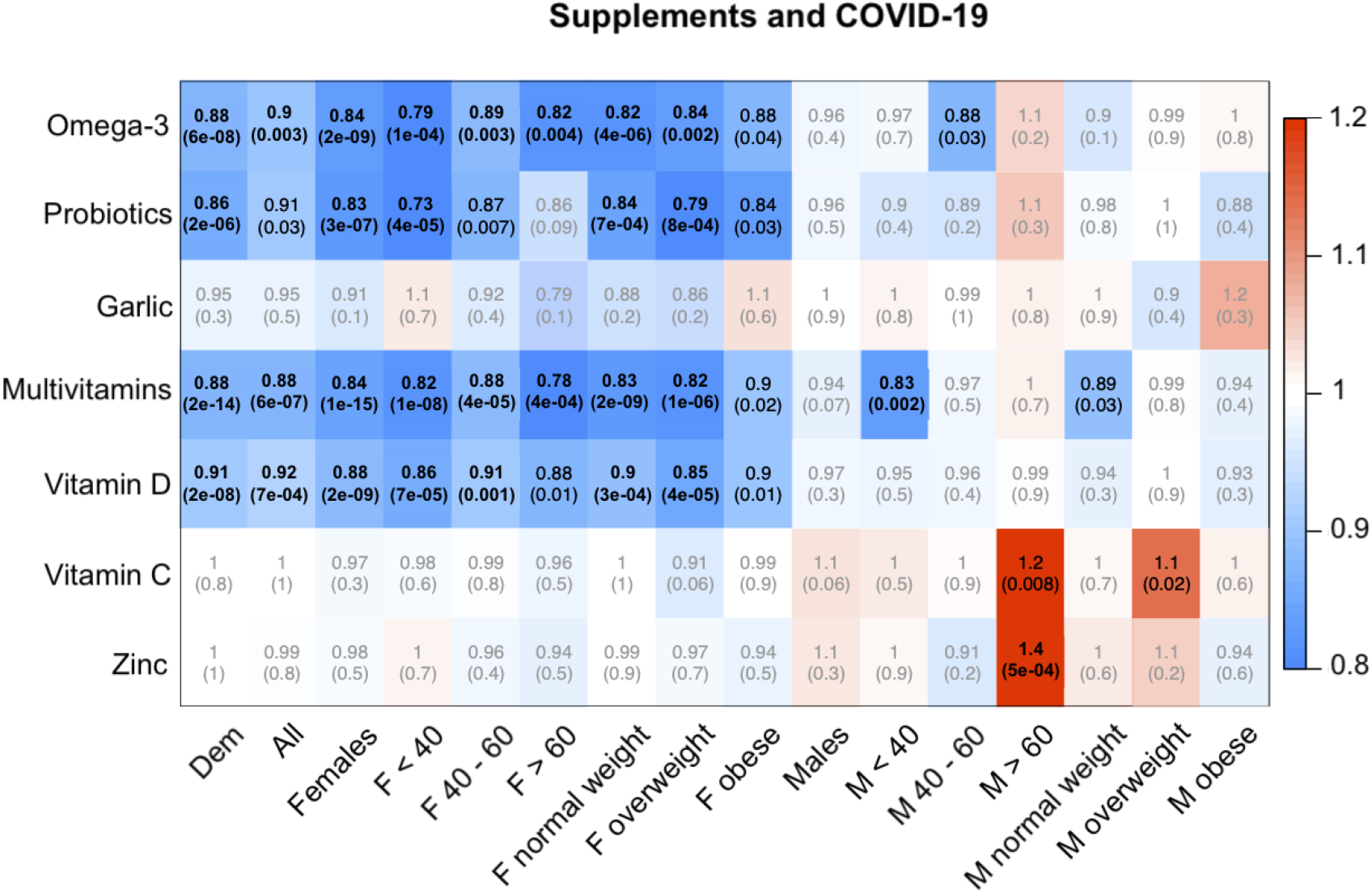
Associations between testing positive for SARS-CoV-2 and self-reported use of supplements in UK app users. Each cell of the matrix displays the odds ratio of the association between use of a type of supplement and testing positive with the corresponding P value in parentheses. The table is colour coded according to the odds ratio, with blue denoting a reduced risk and red denoting an increased risk of testing positive. Bold entries are statistically significant after accounting for multiple testing using Bonferroni correction. Dem = adjusted for age, sex, BMI and health status at sign up; All = adjusted for Dem, index of multiple deprivation, ethnicity, comorbidities (type-2 diabetes, cancer, asthma, heart disease, eczema, hay fever, kidney disease and lung disease), smoking, diet quality; stratified analyses are adjusted for age, (BMI) and health status at sign up as appropriate.

Next, we ran the analyses stratifying by sex, age group and BMI categories and we detected sexual dimorphism in the association between supplement use and testing positive for SARS-CoV-2 (**Figure 1**). We observed a significant protective association of supplement use in females, across all age groups and BMI categories for probiotics, omega-3 fatty acids, multivitamins and vitamin D (OR[95%CI] ranging from 0.73[0.63,0.85] for probiotics in women < 40 years of age to 0.91[0.86,0.96] for vitamin D in women aged between 40-60 years). No protective association was observed in males overall, though in post-hoc subgroup analyses a slight protective effect for multivitamins was observed in men aged < 40 years and in normal weight men and for omega-3 fatty acids in men aged 40-60 years (**Figure 1**). In contrast, there was a positive association in men aged > 60 years and use of zinc supplements (1.4[1.16,1.69], P=4.92×10^−4^) or vitamin C supplements (1.22[1.05,1.41], P=8.1×10^−3^) (**Figure 1**) for testing positive for SARS-CoV-2.

To replicate significant findings from the UK cohort we next used data from the 45,757 US and 27,373 Swedish app users. Cohorts were similar, in that they were also predominantly female (US: 67.8%, SE: 68.6%) and a greater proportion were overweight (BMI(SD), US: 27.3(5.9) kg/m^2^, SE: 26(4.7) kg/m^2^). Overall UK findings were mirrored in both cohorts (**Figure 2**). However, findings by gender were different in different cohorts (**Figure 2**). Associations in females were replicated in the US cohort, but Omega-3 was not associated in Swedish females (**Figure 2**). In males of the US cohort, use of probiotics or vitamin D was associated with decreased risk of a positive test for SARS-CoV-2 (**Figure 2**), while in SE use of probiotics, omega-3 fatty acids, multivitamins or vitamin D was associated with decreased risk of a positive test for SARS-CoV-2 in males (**Figure 2**).

**Figure 2.**
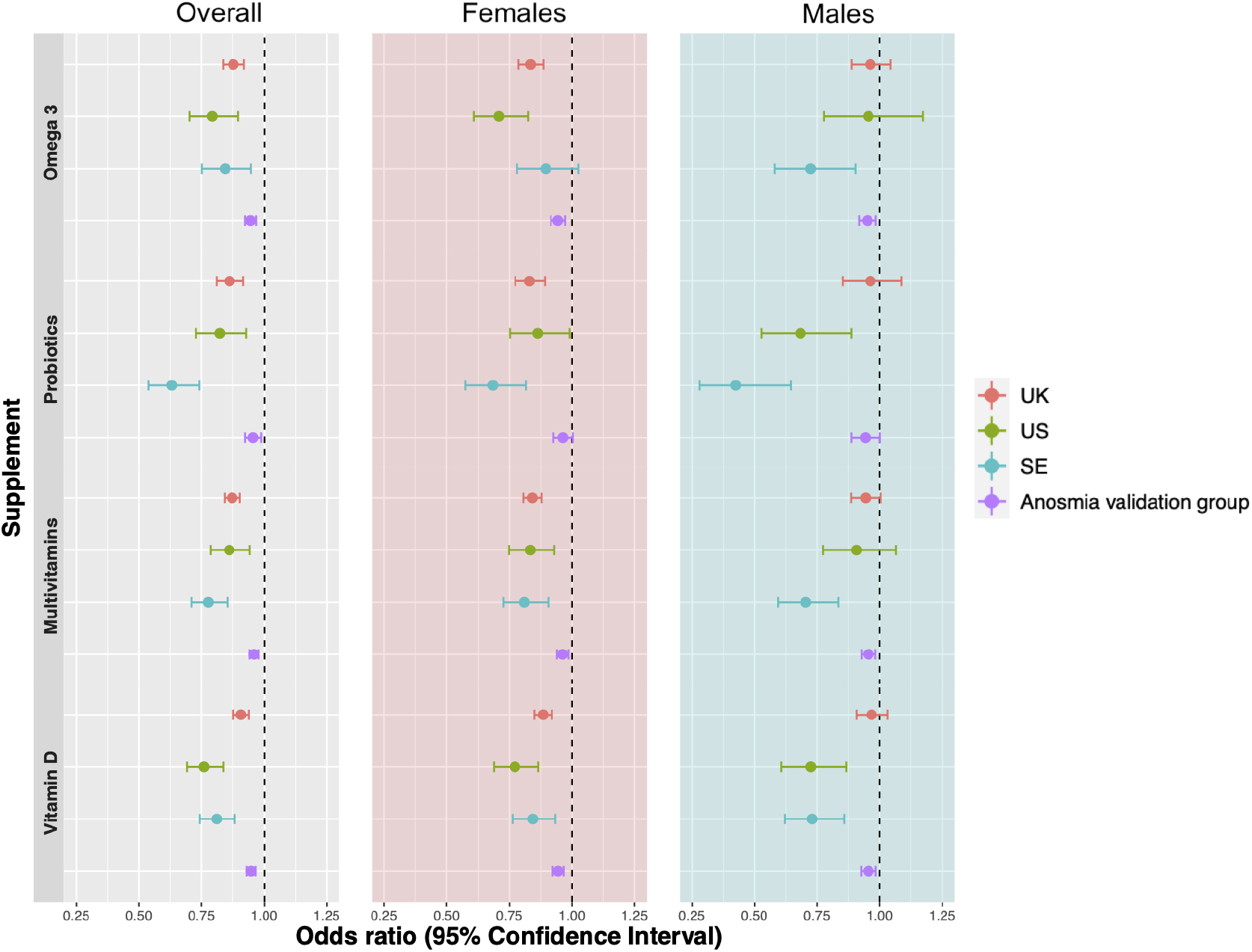
Odds ratios and 95% confidence intervals for the associations between (i) testing positive for SARS-CoV-2 and self-reported use of supplements in three cohorts 372,720 UK, 45,575 US and 27,373 SE, (ii) having new onset anosmia and self-reported use of supplements in 993,365 independent app users. Overall sample analyses are adjusted for age, sex, BMI, and health status at sign up. Analyses according to sex are adjusted for age, BMI and health status at sign up.

Our results were further confirmed in an independent sub-analysis of 993,365 app users not tested for SARS-CoV2 including anosmia cases. We found a small but significant protective effect of around 5% for omega-3 fatty acids, multivitamins, vitamin D and to a lesser extent for probiotics overall and by gender (**Figure 2**).

## Discussion

In the largest observational study on SARS-CoV-2 infection and dietary supplement use to date on over 1.4M app users from three different countries, we show a significant association between use of omega-3 fatty acids, probiotics, multivitamin and vitamin D supplements and lower risk of testing positive for infection with SARS-CoV-2 and developing “COVID anosmia”. However, our stratified analysis in the tested group shows a strong sexual dimorphism with the consistent protective effect present only in females, at least in the UK and for some supplements in the US and SE. This association has several potential explanations including (i) Biological explanations include the well documented discordant immune systems between sexes that could respond differently to supplements [21]. Research indicates that females typically possess a more resilient immune system than males with higher numbers of circulating B cells when matched for age, BMI and clinical parameters [22], as well as a slower age related decline in circulating T-and B-cells [22]. So, it is plausible that supplements could better support the immune system of females than males, although the lack of consistency between countries is problematic as is the lack of gender effect in the larger untested anosmia group; (ii) residual confounding due to behavioural differences between users and non-users or in particular males and females towards infection prevention. Females who purchase vitamins may be more health conscious than males such as greater use of wearing face masks and hand-washing [23– 25]. Indeed, in our data, we found that women tended to wear masks more often than males (44% of women report wearing a mask at least some of the time when outside, compared to 36% of men, P<0.001).

### Vitamin D

A potential antimicrobial role of vitamin D in infections dates back almost a century [26]. Immune cells express the vitamin D receptor and some can synthesise the active form of vitamin D. Vitamin D influences the function of antigen-presenting cells, T-cells and B-cells [27]. It also promotes production of cathelicidin, a microbicidal component of the innate immune system [28]. The overlap between risk factors for vitamin D deficiency and risk of severe COVID-19, such as obesity, age, and ethnicity, gives some plausibility to a protective role of vitamin D [26]. Meta-analysis of a large number, 39, randomised controlled trials identifies that vitamin D modestly reduces the risk of respiratory infections by around 11% with considerable heterogeneity [29]. A Mendelian Randomisation study, on the other hand, suggests that genetic levels of vitamins D are not associated to COVID-19 susceptibility [30], in line with recent results from the UKBiobank[31].

In our data, we find a modest protective effect for infection, with a 9% reduction in risk of testing positive for SARS-CoV-2 in the overall UK cohort, 24% in the US cohort and 19% in the SE group.

### Multivitamins

Multivitamin supplements typically include not just multiple vitamins, but also multiple minerals including trace elements [20]; many of these have antioxidant properties and roles in supporting the immune system [1,2]. Specific micronutrient deficiencies, including of zinc, selenium, vitamin A, vitamin D and vitamin E, have been shown to be detrimental during viral infections [33–35]. Although, some randomised controlled trials have shown that multivitamin supplements reduce the risk of respiratory infections [36], a recent review argues that this evidence is weak and unclear [37]. Here, we provide evidence to support a modest protective effect of multivitamin supplements similar to vitamin D with a 13% reduction in risk of testing positive for SARS-CoV-2 in the overall UK cohort, 12% in the US cohort and 22% in the SE cohort.

### Omega-3 fatty acids

Omega-3 fatty acids can influence antigen presenting cell, T-cell and B-cell function, although their effects on these cell types in humans is not consistently reported. However, they are clearly demonstrated to be anti-inflammatory [38] and to be converted to specialised pro-resolving mediators such as resolvins, protectins and maresins [39]. Whether this is a mechanism by which they reduce risk of testing positive for SARS-CoV-2 is not clear. Nevertheless, here we provide evidence to support a protective effect of omega-3 fatty acid supplements with a 12% reduction in risk of testing positive for SARS-CoV-2 in the overall UK cohort, 21% in the US cohort and 16% in the SE cohort.

### Probiotics

Probiotics modify the host’s gut microbiota and may generate anti-viral metabolites, and they interact with the host’s gut associated immune system [40]. This can result in improved immunity, including enhanced responses to the seasonal influenza vaccine [41]. There is evidence of a gut-lung axis [42], whereby immune effects of microbiota at the gut level can be transferred to the lung, most likely through movement of immune cells. This could explain why some probiotic organisms reduce risk [43–45] and severity [46] of respiratory tract infections. Here we provide evidence to support a modest protective role of probiotic supplements with a 14% reduction in risk of testing positive for SARS-CoV-2 in the overall UK cohort, 18% in the US cohort and 37% in the SE cohort. Effects of probiotics are strain and species specific and we have no information of which probiotics or their quality were being used by participants in this study. Moreover, when we adjusted for other covariates including diet, the effect of probiotics was weaker suggesting that it may be confounded by healthy diet.

### Zinc, Vitamin C and garlic

We saw no protective effects of zinc, garlic or vitamin C. Both zinc and vitamin C have been previously suggested to support the immune system and to prevent respiratory infections <u>[1–30]</u>. Although, their efficacy and evidence base has been questioned and a meta-analysis of vit C showed no preventive benefit and a modest reduction in symptom duration by less than a day [48,49].

Strengths of our study include its large sample size, the confirmation of SARS-CoV-2 through a RT-PCR-or serology-based test, and the replication of the key findings from the UK cohort in two other cohorts, one in the US and the other in SE. Also, our results are broadly replicated when using anosmia as an alternative case definition. Furthermore, we had information on diet quality, and we were able to adjust for it.

Our study also has a number of limitations. First, we used self-reported data which can introduce information bias, including misclassification, or effect bias exposure if they started taking supplements after developing symptoms. We believe this is possible-although would have reduced any real effect. Second, participants using the app were a self-selected group and may not be fully representative of the general population, and may be affected by collider bias [50]. Third, the SARS-CoV-2 infection diagnosis was mainly based on the RT-PCR test that has less than 100% sensitivity (true positive rate) [51,52]. Fourth, participants might have been taking supplements in addition to the seven we asked them about. Fifth, we do not know the exact intakes of the ingredients within the supplements used by the participants. Finally, this is an observational cross-sectional study captured during a specific timeframe, and our study design does not allow an inference of causality.

In conclusion, our data find a correlation between use of multivitamins, omega-3 fatty acids, vitamin D and probiotics and slightly lower risk of SARS-CoV-2 infection in women in the UK, US and SE, but no effect of zinc, vitamin C or garlic. The larger anosmia data confirmed a more modest effect. Given the interest in supplements during the pandemic, large randomised controlled trials of selected supplements testing their protective effects and also possible adverse effects on disease severity are required before any evidence-based recommendations can be made. We eagerly await the result of ongoing trials, including vitamin D and COVID risk [53,54].

### Section 1: What is already known on this topic

- Dietary supplements have been shown to play key roles in supporting immune function, but the extent to which specific supplements are associated with reduced risk of SARS-CoV-2 infection is not known.

### Section 2: What this study adds

- Individuals taking multivitamins, omega-3 fatty acids, probiotics or vitamin D were less likely to be tested positive for SARS-CoV-2 in three large independent cohorts of app users.
- There was a significant protective association for vitamin D, omega-3 fatty acids, probiotics and multivitamins in females across all ages and BMI categories within the largest (UK) cohort. Yet, there was no association in men of this cohort.
- Vitamin C, zinc and garlic supplements had no association with risk for SARS-CoV-2.
- There is a need for randomised controlled trials of selected supplements.

## Acknowledgements

We express our sincere thanks to all the participant users of the app, including study volunteers enrolled in cohorts within the Coronavirus Pandemic Epidemiology (COPE) consortium. We thank the staff of Zoe Global Limited, the Department of Twin Research at King’s College London, the Clinical & Translational Epidemiology Unit at Massachusetts General Hospital, Researchers and staff at Lund University in Sweden for their tireless work in contributing to the running of the study and data collection.

Zoe provided in kind support for all aspects of building, running and supporting the app and service to all users worldwide. The Department of Twin Research is funded by the Wellcome Trust, Medical Research Council, European Union, Chronic Disease Research Foundation (CDRF), Zoe Global Ltd and the National Institute for Health Research (NIHR)-funded BioResource, Clinical Research Facility and Biomedical Research Centre based at Guy’s and St Thomas’ NHS Foundation Trust in partnership with King’s College London. CM is funded by the Chronic Disease Research Foundation and by the MRC Aim-Hy project grant. PL is founded by the CDRF, SO is funded by the Wellcome/EPSRC Centre for Medical Engineering (WT203148/Z/16/Z), Wellcome Flagship Programme (WT213038/Z/18/Z), and PCC is supported by the National Institute for Health Research Southampton Biomedical Research Centre.

We express our sincere thanks to all the participants of the *COVID Symptom Study* app. We thank the staff of Zoe Global Limited and the Department of Twin Research for their tireless work in contributing to the running of the study and data collection.

---

**Table S1.**
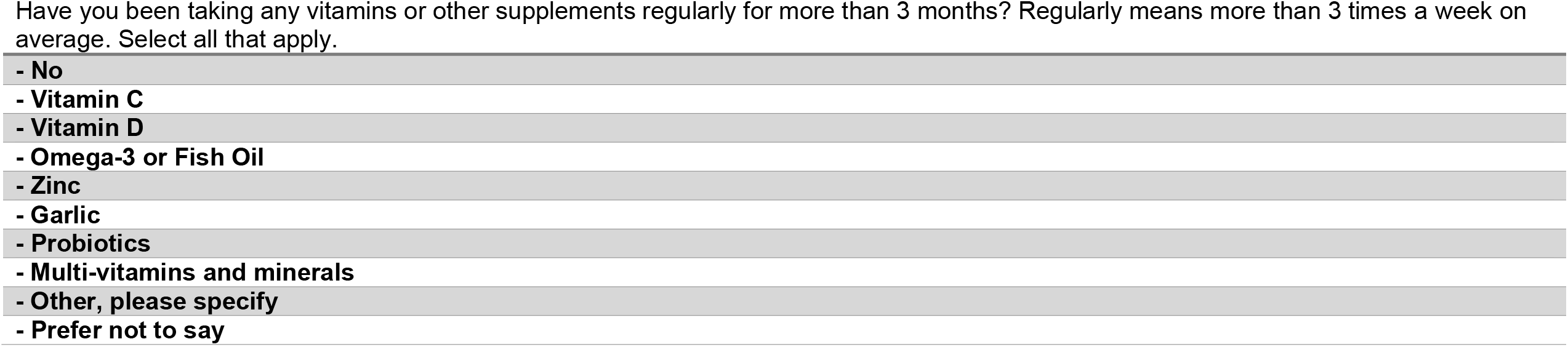
List of questions on supplements usage.

|  |
| --- |
| - No |
| - Vitamin C |
| - Vitamin D |
| - Omega-3 or Fish Oil |
| - Zinc |
| - Garlic |
| - Probiotics |
| - Multi-vitamins and minerals |
| - Other, please specify |
| - Prefer not to say |

